# Generalizability in real-world trials

**DOI:** 10.1101/2024.01.10.24301117

**Authors:** Anatol-Fiete Näher, Marvin Kopka, Felix Balzer, Matthias Schulte-Althoff

## Abstract

Real-world evidence (RWE) trials have a key advantage over conventional randomized controlled trials (RCTs) due to their possibly higher external validity. This allows for better generalizability of results to larger populations, which is essential for evidence-based decision making in clinical medicine, pharmacoepidemiology, and health policy. Random sampling of RWE trial participants is regarded the gold standard for generalizability. Additionally, the use of sample correction procedures can increase the generalizability of trial results, even when using non-randomly sampled real-world data (RWD). This study presents descriptive evidence on the extent to which the design of currently planned or already conducted RWD/E trials takes sampling into account. It also examines whether random sampling or procedures for correcting non-random samples are considered. Based on text-mining of publicly available metadata provided during registrations of RWD/E trials on clinicaltrials.gov, EU-PAS, and the OSF-RWE registry, it is shown that the share of RWD/E trial registrations with information on sampling increased from 65.27% in 2002 to 97.43% in 2022, with a corresponding increase from 14.79% to 28.30% for trials with random samples. For RWD/E trials with non-random samples, there is an increase from 0.00% to 0.22% of trials in which sample correction procedures are used. We conclude that the potential benefits of RWD in terms of generalizing trial results are not yet being fully realized.

## Introduction

According to the US Food & Drug Administration (FDA), real-world data (RWD) are “data relating to patient health status and/or the delivery of health care routinely collected from a variety of sources”.^1^ The sources from which RWD can be obtained range from disease registries, insurance claims and population surveys to wearables, apps and electronic health records.^2,3^ Trials based on RWD, i.e. Real-World Evidence (RWE) trials, are more efficient to conduct, can include investigations of outcomes that would not be possible with RCTs, and include observations of real-world patient behavior outside the setting of common clinical trials.^4^ However, a key advantage of RWD/E trials is that they offer greater external validity than RCTs.^5-8^ The results from RWD/E trials are potentially generalizable to broader populations. The basic requirement for such generalizability is random sampling of study participants: random sampling involves specifying a probability *p ∈ (0,1)* for each study participant to be included in the trial sample,^3,9^ thereby avoiding selection bias. Selection bias implies that outcomes from real-world trials cannot be unreservedly generalized to all population groups of interest. One reason for this is that selection bias distorts statistical estimates of treatment effects. A second reason is that most inferential statistical methods used in real-world studies are based on normality assumptions, which are only met with random sampling.

Hence, in principle, sampling methods must be distinguished according to whether they are suitable for generating random samples or not. Non-random sampling per se does not preclude the use of RWD. On the one hand, the generalizability of results does not necessarily have to be the goal of every RWD/E trial. For instance, the primary objective may be to record patient behavior in order to obtain more detailed information about the effects of a tested drug. If, on the other hand, the generalizability of results is one of the objectives of an RWD/E study, it must be assumed that in many cases random samples will not be available or can only be obtained at great cost and in a time-consuming manner. This is particularly true for trials with rare disease endpoints, as the cost and time required to obtain RWD increases as the disease prevalence decreases. In such cases where random samples are not available, correction procedures can be used. Weighting or raking^10-12^ as well as sample selection^13-15^ and outcome regression models^16,17^ can help to improve the generalizability of results from non-random samples in RWD/E studies.^3,9^

Regarding the generalizability of findings from RWD/E trials, there is currently a lack of transparency. It is worth noting that both the FDA and the European Medicines Agency (EMA) have established regulatory pathways for RWE,^18-21^ which determine the design of RWD/E trials. In principle, standardized study protocols can be used to increase transparency along these pathways regarding the generalizability of findings from RWD/E studies. However, existing protocols, such as the Harmonized Protocol Template to Enhance Reproducibility (HARPER), do neither document random or non-random sampling nor the use of sample correction procedures.^22^ Information from current trial protocols does not allow conclusions to be drawn about the generalizability of results.

Also, trial registration can increase the transparency of RWD/E trials regarding the generalizability of findings. In 2019, the ISPO-ISPER working group proposed registrations of RWD/E trials as part of its Real-World Evidence Transparency Initiative.^23^ As suggested by ISPO-ISPER, trial registration involves documenting the entire research design in order to track deviations from standardized study designs and to improve the overall transparency of individual trials.^23^ Currently, there are three repositories available for registering RWD/E trials. The first is the OSF Real-World Evidence (OSF-RWE) Registry, which is offered by ISPOR in conjunction with the Open Science Framework. The second is the European Union’s electronic registry for post-authorization studies (EU-PAS), which is frequently used for Phase IV trial registration. Finally, clinicaltrials.gov, originally developed for clinical trials, also permits the registration of RWD/E trials. In terms of transparency, a key advantage of trial registries over standard protocols is that trial design information can be provided voluntarily at the time of registration, going beyond what is required in existing standard protocols. In fact, clinicaltrials.gov registrations even require information on random or non-random sampling. In addition, other information, such as information on the use of sample correction procedures, can be provided on a voluntary basis, although providing this information is not mandatory for registration in any of the clinicaltrials.gov, OSF-RWE and EU-PAS repositories.

On the basis of information that may be contained in publicly available trial registration documents, it is therefore possible to assess the extent to which considerations of random or non-random sampling and, where appropriate, sample correction procedures in the design of registered RWD/E trials are presented transparently. The transparent presentation of sampling procedures or procedures for the correction of non-random sampling is a prerequisite for the assessment of the generalizability of findings from RWD/E trials. This study provides descriptive evidence regarding the extent to which information on random and non-sampling has been shared to date in registrations of RWD/E trials on clinicaltrials.gov, OSF and EU-PAS. For trials with non-random sampling on clinicaltrials.gov, OSF and EU-PAS, we also report the extent to which the utilization of sample correction procedures is reported. Our study thus contributes to increasing transparency regarding the generalizability of results from RWD/E trials.

## Methods

Our descriptive study first describes the number of registrations of RWD/E trials on clinicaltrials.gov and in the OSF and EU-PAS registries. In order to report how many of the RWD/E trials listed in these databases provided information on random and non-random sampling as well as sample correction procedures, we first extracted publicly available study metadata from all three databases, where available. Trial submissions were eligible for inclusion if they were explicitly designed to utilize RWD in a non-interventional trial setting. The number of RWD/E registrations was calculated based on the number of trials registered as observational studies in clinicaltrials.gov, OSF and EU-PAS. To gather information on the number of trials providing information on random and non-random sampling and sample correction procedures, the metadata obtained for each registry were searched separately using natural language processing. We utilized a keyword search algorithm that allowed us to capture text in all database fields. The same search strings were applied to all three databases. The search was confined to registrations in English language dating back to the earliest ones in all three databases. Hence, the period between 2002 for clinicaltrials.gov, from 2009 for EU-PAS and from 2021 for OSF to 2022 is reported. The database searches were conducted in November 2023 and were not subsequently updated. A table of the elements of the defined search strings can be found in the appendix.

### Limitations

This study has several limitations. First, despite a thorough search, registries other than clinicaltrials.org, EU-PAS and OSF may have been missed. This may have led to the omission of some publicly available registered RWD/E trials. In addition, due to the large number of records identified, not all trial metadata could be screened manually. As a result, some descriptions in the text fields may not have been considered. For instance, clinicaltrials.org not only requires the reporting of sampling methods, but also provides a specific field for this information. In contrast, OSF and EU-PAS registries do not have a specific field for sampling methods. For this reason, researchers can only deliberately report sampling methods used in open text fields for trial descriptions. Due to the use of natural language processing to extract information, we may have overlooked some trials where sampling approaches were described using different terminologies. However, the search was carried out after the selection of keywords had been reviewed by a larger group of scientific peers. Thus, the risk of omission is considered low. Third, the ability or need to generalize the results of RWD/E trials arises only if the study population represents a subset of the population of interest. If the study population represents the entire population of interest, the results of the study cannot or need not be generalized. Thus, the results of a share of those trials for which sampling methods are not reported may apply to the entire population of interest. In these cases, there is no possibility or need to generalize results. We assume that the share of trials in question is small, because in the vast majority of trials a complete survey of all individuals in a population of interest is not feasible.^9,24^

## Results

A total of 455,081 registrations were identified over the period from 2002 to 2022. Of these, 452,752 (99.49 %) were registered on clinicaltrials.gov and 2,275 (0.5 %) on the EU-PAS and 54 (0.01 %) on the OSF databases. As illustrated in Figure 1, the total yearly number of registered RWD/E trials increased from 622 in 2002 to 7,939 in 2022. This represents a compound annual increase of 13.14%. The proportion of RWD/E trials in all registered trials has been on the rise from 15.12% (2002) to 24.19% (2022).

**Figure 1.**
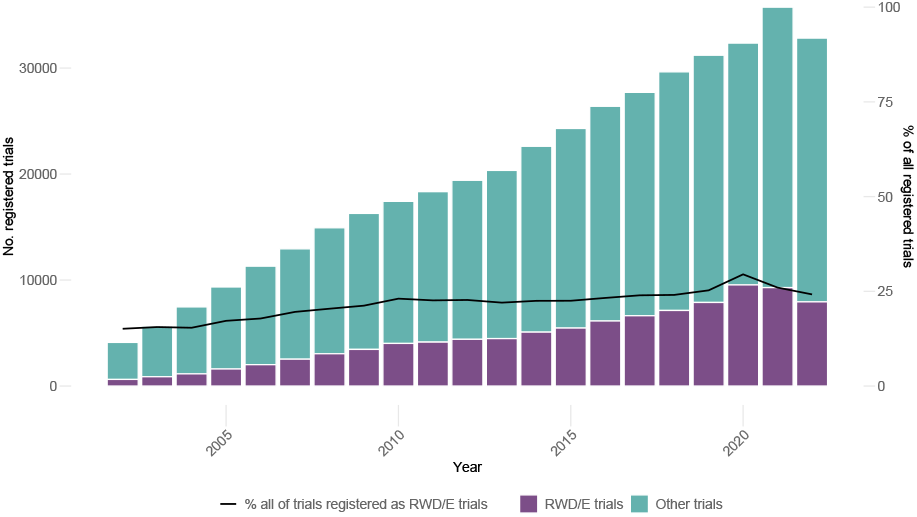
Absolute numbers and shares of RWD/E trials among all registered trials. Sources: clinicaltrials.gov, OSF-RWE & EU-PAS registries.

Similarly, as depicted in Figure 2, shares of registered RWD/E trials providing information on the sampling methods used in all RWD/E studies increased from 65.27% in 2002 to 97.43% in 2022. Furthermore, the yearly numbers of registered RWD/E studies that utilized random sampling have increased from 92 in 2002 to 2247 in 2022, representing an average annual growth rate of 17.33%. Figure 2 also demonstrates that shares of RWD/E studies with random sampling in all RWD/E trials has remained stable from 2007 to 2022: percentages of RWD/E trials with random sampling increased from 14.79% to 25.31% in 2007, with less significant increases in the following years until 2022 (28.30%).

**Figure 2.**
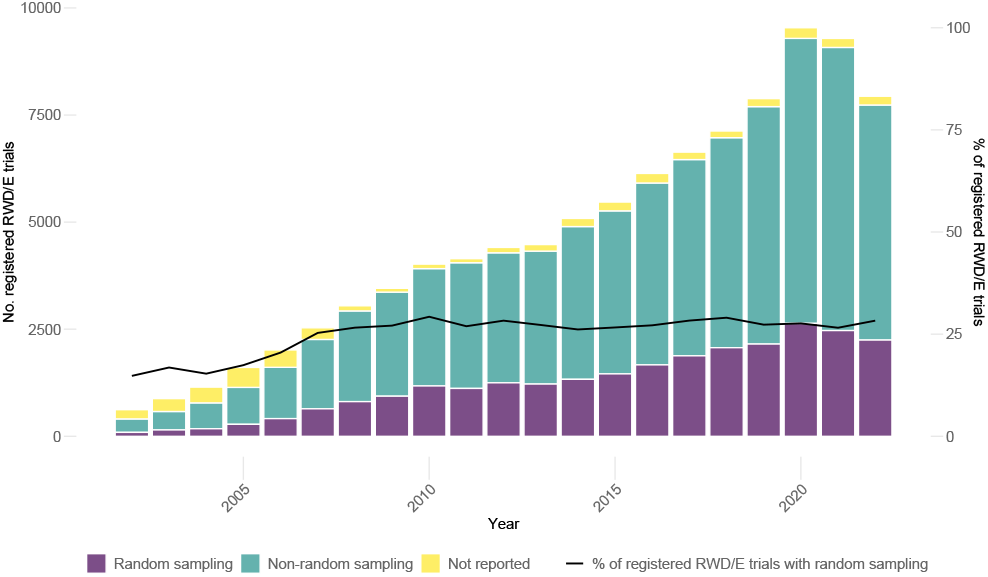
Shares of trials reporting random or non-random sampling among all registered RWD/E trials. Sources: clinicaltrials.gov, OSF-RWE & EU-PAS registries.

Figure 3 illustrates the increase in RWD/E studies with non-random sampling from 2002 to 2022. A visual inspection of the lower panel of figure 3 reveals that the number of RWD/E trials with non-random sampling, in which sample correction procedures were planned, grew from 2 in 2006 to 12 in 2022, with an average annual growth rate of 11.85%. However, as the lower panel of Figure 3 also shows, shares of sample correction procedures applied in RWD/E trials with non-random sampling remained consistently low over the entire observation period: between 2002 and 2022, an increase from 0.00 % to 0.22 % was observed.

**Figure 3.**
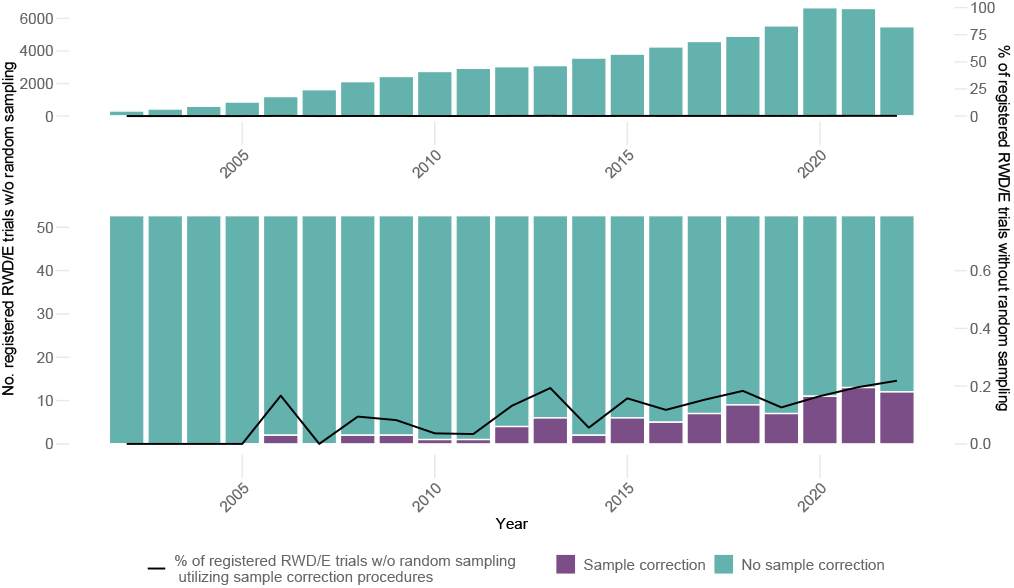
Shares of trials reporting the (planned) use sample correction procedures among all registered RWD/E trials with non-random samples. The lower panel illustrates a larger scale of the upper panel in order to depict the small shares of trials with sample correction methods. Sources: clinicaltrials.gov, OSF-RWE & EU-PAS registries.

Table 1 presents results categorized by trial registry. The shown results are aggregated over the entire observation period. It is evident that clinicaltrials.gov accounts for the largest share of registered RWD/E trials (97.61%), followed by EU-PAS (2.32%) and OSF (0.06%). Additionally, the share of registrations of RWD/E trials with no information on the sampling methods used is lowest on clinicaltrials.gov (4.02% vs. 96.62% on EU-PAS and 88.89% on OSF). The share of RWD/E studies that provide information on the use of random sampling methods is highest on clinicaltrials.gov (26.90%), compared to EU-PAS (3.34%) and OSF (11.10%). Similar findings are observed in RWD/E trials with non-random sampling (60.08% on clinicaltrials.gov vs. <0.01% on EU-PAS and 0.00% on OSF). In contrast, shares of RWD/E trials with non-random sampling and sample correction procedures remain consistently low across all three registries. On clinicaltrials.gov the share amounts to 0.13%, while there are no shares on EU-PAS and OSF (0.00% and 0.00%).

**Table 1.** Shares of trials reporting random or non-random sampling and sample correction procedures among registered all registered RWD/E trials. Sources: clinicaltrials.gov, OSF-RWE & EU-PAS registries.

|  | clinicaltrials.gov | EU-PAS | OSF |
| --- | --- | --- | --- |
| Number of RWD/E trials among all RWD/E trial registrations, n (%) | 102,471 (97.77%) | 2,275 (2.17%) | 54 (0.05%) 97.645 |
| RWD trials reporting sampling procedures among all RWD/E trial registrations, n (%) |  |  |  |
| Random Sample | 27.562 (26.90%) | 76 (3.34%) | 6 (11.10%) |
| Non-random sample | 70.787 (69.08%) | 1 (<0,01%) | 0 (0%) |
| Not reported | 4.122 (4.02%) | 2.198 (96.62%) | 48 (88.89%) |
| RWD/E with sample correction procedures among all registered non-random trials, n (%) | 90 (0.13%) | 0 (0.0%) | 0 (0.0%) |

## Discussion

Over the observation period from 2002 to 2022, the share of RWD/E studies in all clinical trials has risen continuously and was just under a quarter during the last observation year. Thus, it can be assumed that some of the advantages of RWD over RCT data, such as information on health-related behavior or patient-reported outcomes, are used on a regular basis in the context of registered trials. However, this does not apply in the same extent to the generalizability of trial results, which is one of the key advantages of RWD: on the one hand, the share of registered RWD/E trials reporting on the sampling method employed also increased between 2002 and 2022 and was approaching 100% in 2022. On the other hand, this trend differs significantly depending on the registry considered: the rise in reported sampling methods is primarily due to clinicaltrials.gov’s requirement for trial registrations to include information on the sampling method used. The absence of sampling information in 4.02% of RWD/E trials registered on clinicaltrials.gov is due to registrations made before the requirement to provide such information. In contrast, the share of RWD/E trials without information on sampling methods is significantly higher in EU-PAS and OSF at 96.62% and 88.89%, respectively. In neither of these registers is the provision of information on sampling methods a prerequisite for registration.

When registering RWD/E trials on clinicaltrials.org, it is further necessary to specify whether the trial to be registered is based on random sampling or whether the data are generated by non-random processes. Compared to trials registered on EU-PAS and OSF, clinicaltrials.org has a higher share of RWD/E trials with random sampling (26.90% vs. 3.34% and 11.10%). At a comparatively low level, a similar pattern can be observed for the use of sample correction procedures in non-randomized RWD/E trials. The increase to 0.22% in 2022 results from the fact that sample correction procedures have so far only been used or planned for RWD/E trials with non-random sampling registered on clinicaltrials.org (0.13% vs. 0.00% and 0.00%). One explanation for the higher share of RWD/E trials with generalizable results on clinicaltrials.org may be that the mandatory information on sampling methods leads to an increased awareness among researchers of the limited generalizability associated with non-random samples. This may, in turn, lead to greater attention to the generalizability of results in the design and conduct of more RWD/E trials. This would provide an approach for a cross-registry policy to improve the generalizability of RWD/E trials. However, these hypotheses cannot be tested with the data available for this study. Data suitable for testing the hypotheses should be collected in future prospective studies.

Currently, just under a third of registered RWD/E studies use either sampling or sampling correction methods in order to increase generalizability. This means that one of the great potentials of RWD remains untapped in the majority of cases. As mentioned at the beginning, this may also be due to the fact that RWD is not necessarily used with the primary aim of increasing generalizability. However, current calls to improve generalizability^25^ refer to all forms of RWD/E trials in addition to RCTs. Thus, when conducting RWD/E studies, random sampling should be used more than in the past or, if this is not possible, sample correction procedures should be applied. To this end, when registering RWD/E trials, registries and regulatory authorities should take measures to ensure the highest possible generalizability across registries. Further measures should also be taken to verify that the chosen approaches for sampling or sample correction are the most appropriate for specific trial demands. In addition to mandatory reporting of sampling procedures, the information provided as part of trial registrations could be subject to peer review with a checklist including details of the sampling strategy. Based on this procedure, the selection of a different sampling strategy or adjustments for non-probability samples could be recommended, if necessary. By defining appropriate procedures at an early stage, the generalizability of results in RWE/D studies can be significantly improved.

Overall, there is room for improvement in the generalizability of RWD/E trials. It should be noted, however, that randomized selection of study participants is often difficult to implement in RWD/E trials. Above policy measures should therefore be further complemented by trust-building measures that increase participation in trials and the willingness to share health data. Moreover, access to registries and claims data should also be facilitated in order to simplify the definition of reference populations for sampling or the well-guided selection of sample correction procedures. Widespread implementation of these measures will ensure that therapies are better adapted to the needs of patient groups that are currently under-represented in trials, thereby leading to further improvements in evidence-based care.

## Data Availability

All data produced in the present study are available upon reasonable request to the authors

### Appendix

**Table A1.** Concepts and search strings used in text mining analysis.

| Concept | Search string elements |
| --- | --- |
| Random Sampling | • Random sample<br>• Sampled randomly<br>• Randomly chosen<br>• Chosen randomly<br>• Sample randomly<br>• Stratified sampling<br>• Systematic sampling<br>• Systematically chosen<br>• Chosen systematically<br>• Sampled systematically<br>• Sample systematically<br>• Cluster sampling<br>• Sampled in clusters<br>• Sample in clusters |
| Non-Random Sampling | • Non-probability sample<br>• Non-random sample<br>• Convenience-based sample<br>• Convenience-based sampling<br>• Convenience sampling<br>• Convenience sample<br>• Judgmental sample<br>• Judgment-based sample<br>• Judgment-based sampling<br>• Judgmental sampling<br>• Sampled judgmentally<br>• Sample judgmentally<br>• Purposive sample<br>• Purposive sampling<br>• Sampled purposively<br>• Sample purposively<br>• Snowball sample<br>• Quota sample<br>• Volunteer sample<br>• Sample of volunteers<br>• Volunteer sampling<br>• Self-selected sample<br>• Self-selected sampling<br>• Self-selected sample<br>• Expertise-based sampling<br>• Expertise-based sample<br>• Expert sample<br>• Expert-based sample\|<br>• Expert-based sampling |
| Sample Correction Procedure | • Hackman<br>• Vella<br>• Heckman correction<br>• Self-selection model<br>• Self selection model<br>• Selfselection model<br>• Sample selection model<br>• Selection bias model<br>• Survey weight<br>• Raking<br>• Rake<br>• Post-stratum adjustment<br>• Poststratum adjustment<br>• Post stratum adjustment<br>• Stratification adjustment<br>• Post-stratified weight<br>• Poststratified weight<br>• Post-stratified weight<br>• Post stratified weight<br>• Poststratified analysis<br>• Post-stratified analysis<br>• Post stratified analysis<br>• Post-stratified estimation<br>• Post stratified estimation<br>• Poststratified estimation<br>• Stratification correction<br>• Weighting by strata<br>• Stratum-specific weighting<br>• Stratum specific weighting<br>• Stratum-based adjustment<br>• Stratum based adjustment<br>• Post-stratification technique<br>• Full Information Maximum Likelihood<br>• FIML<br>• Poststratification technique<br>• Inverse probability weighting<br>• Survey weighting adjustment<br>• Rim weighting<br>• Selection bias correction<br>• Non-response adjustment<br>• IPTW<br>• Inverse probability of treatment weighting<br>• IPW<br>• Inverse-probability weighting<br>• Inverse probability weighting |

## Notes

### Competing Interest Statement

The authors have declared no competing interest.

### Funding Statement

This study did not receive any funding

